# Comparison of healthcare resource use and cost between influenza and COVID-19 vaccine coadministration and influenza vaccination only

**DOI:** 10.1101/2024.06.06.24308506

**Authors:** Darshan Mehta, Tianyu Sun, Jane Wang, Aaron Situ, Yoonyoung Park

**Author notes:** **Address for correspondence,** Darshan Mehta.

## Abstract

**Objective:** To compare healthcare recourse utilizations (HCRU) and all-cause medical costs among individuals 50 years or older who received both influenza and COVID-19 vaccines on the same day and those that received influenza vaccine only

**Methods:** We conducted a retrospective cohort study leveraging Optum’s de-identified Clinformatics® DataMart from 8/31/2021 to 7/31/2023. Individuals 50 years or older continuously enrolled in health plans for one year prior and until 7/31/2023 were included in the study. Subjects were categorized into 2 cohorts based on their vaccination status between 8/31/2022 and 1/31/2023: receiving influenza and COVID-19 vaccine on same day (co-admin cohort) and receiving influenza vaccine only (influenza cohort). The association between vaccination status and all-cause, influenza-related, COVID-related, pneumonia-related, and cardiorespiratory-related hospitalization, as well as outpatient or emergency room visits and all-cause medical costs was estimated by weighted generalized linear models, adjusting for confounding by stabilized inverse probability of treatment weighting.

**Results:** 613,156 (mean age: 71) and 1,340,011 (mean age: 72) individuals were included in the co-admin and influenza cohort respectively. After weighting, the baseline characteristics were balanced between cohorts. The co-admin cohort was at statistically significant lower risk of all-cause (RR: 0.95, 95%CI: 0.93-0.96), COVID-19-related (RR: 0.59, 95%CI: 0.56-0.63), cardiorespiratory-related (RR: 0.94, 95%CI: 0.93-0.96) and pneumonia-related (RR: 0.86, 95%CI: 0.83-0.90) hospitalization but not influenza-related hospitalizations (RR: 0.91, 95%CI: 0.81, 1.04) compared to influenza cohort. Co-administration was associated with 3% lower all-cause medical cost (cost ratio: 0.974, 95% CI: 0.968, 0.979) during the follow-up period compared to receiving influenza vaccine only.

**Conclusion:** Receiving both COVID-19 and influenza vaccines on the same day in comparison to receipt of influenza vaccine only was associated with reduced risk of HCRU, especially COVID-19 related hospitalization and all-cause medical costs. Interventions that increase vaccine coverage, particularly for COVID-19 might have public health and economic benefits.

## Background

Acute respiratory diseases caused by SARS-CoV-2 and influenza viruses impose a significant health burden. Since the pandemic COVID-19 has resulted in 6.9 million hospitalizations and 1.1 million deaths in the United States (US).^1^ In 2023, CDC estimates there were 850,000 hospitalizations and 70,000 deaths due to COVID-19, most of them among older individuals. In the same year CDC estimates influenza caused up to 790,000 hospitalizations and up to 69,000 deaths.^2^

Vaccines exist to protect the population against both COVID-19 and influenza disease. For the 2023/2024 season, CDC currently recommends influenza and COVID-19 vaccination for everyone greater than 6 months of age ^3,4^ CDC recommends co-administration of both influenza and COVID-19 vaccines at the same visit for eligible individuals if timing is aligned.^5^ Despite the CDC recommendations and prevailing unmet needs, COVID-19 vaccine uptake in the US has declined over time.^6^ In the 2023/24 season, 22.5% of adults and 32.3% individuals with health conditions associated with higher risk of adverse outcomes received the COVID-19 vaccination.^1^ The influenza vaccine coverage estimated by the CDC is relatively higher in 50 to 64 age group at 51.5% and in 65 years and older at 73.8%. ^7^ Ensuring sufficient vaccine coverage for both COVID-19 and influenza is crucial to reduce the associated significant burden of respiratory illnesses.

The suboptimal vaccine coverage leaves many adults and especially high-risk individuals like older adults at increased risk of severe influenza and COVID-19. Given the annual circulation and continuous evolution of both influenza and SARS-CoV-2, there remains a need to improve simultaneous vaccination uptake, especially in the most vulnerable populations, such as the elderly. Co-administration of vaccines during the same visit could be one strategy to increase the vaccination coverage rate.^8^ However, literature is still evolving about the medical and cost outcomes associated with co-administration of the two vaccines in older adults. In this study, we examined the downstream HCRU and medical cost following co-administration of COVID-19 and influenza vaccines on the same day, compared to receiving influenza vaccine only in a given season.

## Methods

### Study design and participants

We conducted the study using the Optum Clinformatics Data Mart (CDM). The dataset is comprised of closed medical and pharmacy claims including a large proportion of Medicare Advantage plans. Data from August 31, 2021, to July 31, 2023, was used for the analyses and data extraction was performed on March 29, 2024. Adults greater than 50 years or older on the index date and continuously enrolled in health plans (no gap allowed) one year prior to index date and up to July 31, 2023, were eligible for the study. Index vaccination status was assessed between August 31, 2022, and January 31, 2023 (identification period), covering majority of the 2022/2023 seasonal vaccination season in the US. We included individuals who received at least one influenza vaccine or an influenza and COVID vaccine on the same day during the identification period. Key exclusion criteria were individuals with missing age, sex, insurance type and region information. We excluded patients who received influenza or COVID-19 vaccine during August 1-30, 2022, to exclude potential prior season vaccination. Individuals who received more than one influenza vaccine or more than one COVID-19 vaccine during August 31, 2022 –July 31, 2023, were excluded (**Figure 1**).

**Figure 1:**
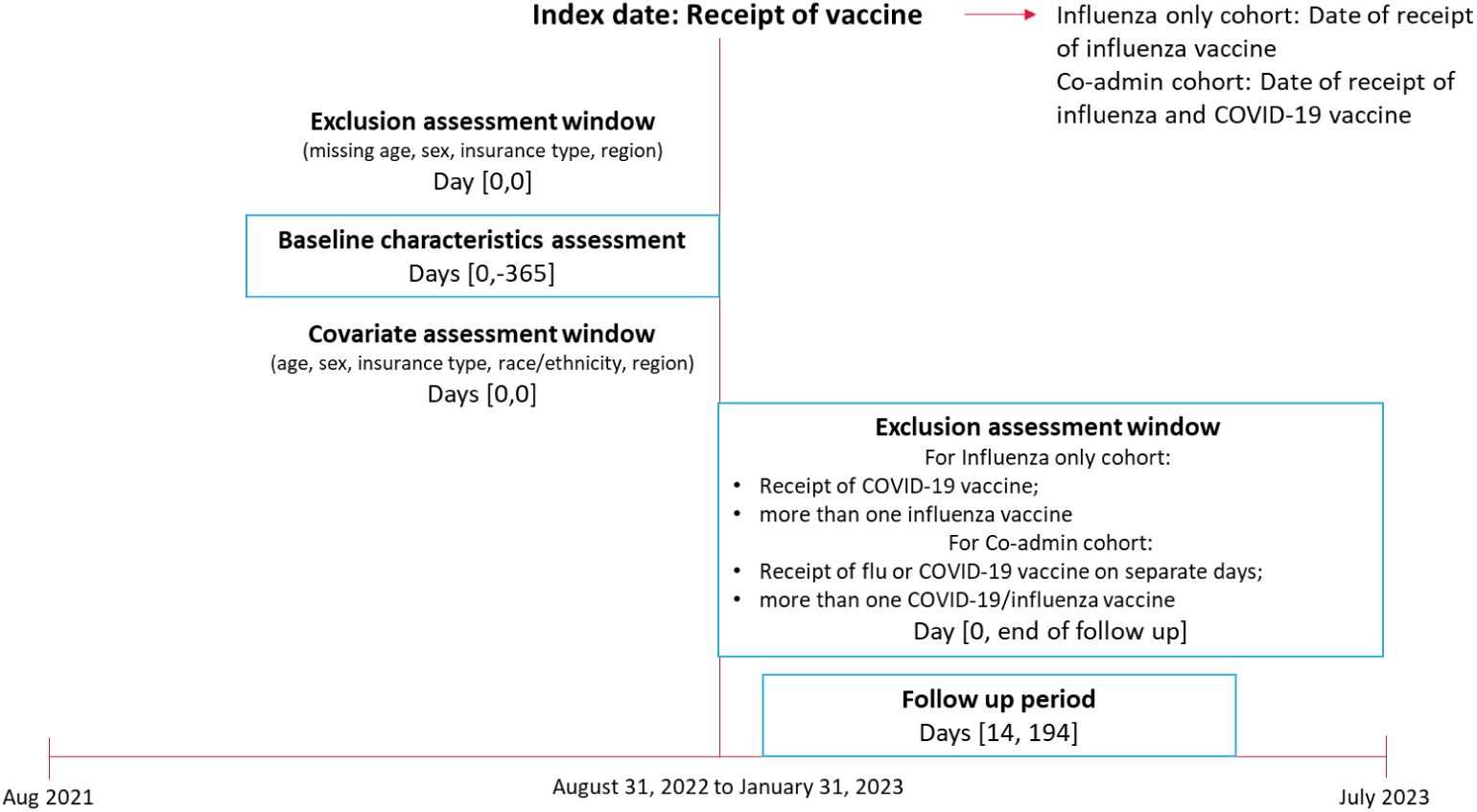
Description of study design.

The population was categorized into 2 cohorts based on their vaccination status during the identification period: those receiving influenza and COVID-19 vaccine on same day (co-admin cohort), aligned with CDC definition of co-administration^5^ and those receiving influenza vaccine only (influenza cohort). For the influenza cohort, individuals who received COVID-19 vaccine between index date and July 31, 2023, were excluded. Similarly, for co-admin cohort, individuals who received influenza and COVID-19 vaccine on separate days were excluded. As the unvaccinated cohort would be considered different in terms of unobserved characteristics than people who got either COVID-19 or flu vaccine, to avoid the risk of unmeasured confounding we did not consider them as our referent.^9^ Further, among individuals who got either COVID-19 or flu vaccine we considered the flu only cohort as our control as flu vaccination coverage rate is higher than COVID-19 vaccine coverage rate.

Study subjects were indexed by their date of vaccination for influenza and COVID-19 (co-admin cohort), or their date of vaccination for influenza (influenza cohort). Baseline period for covariate assessment was defined as 365 days prior to the index date, and follow-up period was defined as from 14 days post-index date to 194 days after the index date (i.e., 6 month of follow-up), up to July 31, 2023.

### Exposure and Outcomes

Vaccination status as the exposure was determined using Current Procedural Terminology (CPT), Healthcare Common Procedure Coding System (HCPCS), and National Drug Codes (NDC) from any care setting (Supplementary Table 1). The healthcare resource utilization (HCRU) outcomes were defined as having one or more all-cause, influenza related, COVID-19 related, pneumonia-related and cardiorespiratory-related hospitalizations during the follow-up, the number of outpatient or emergency room (OP/ER) encounters during the follow-up, and all-cause medical incurred during the follow-up. All-cause HCRU was defined as hospitalizations or OP/ER encounters due to any cause. Influenza, COVID-19, pneumonia-related and cardiorespiratory related HCRU was defined as hospitalization and OP/ER encounters with prespecified ICD-10 diagnosis codes (Supplementary Table 2) in any position associated with the claim. All-cause medical cost was defined as the medical cost associated with hospitalization or OP /ER counters due to any cause. The mean cost per patient was determined over the follow-up period. Costs were reported in 2023 USD.

### Covariates

Demographic characteristics such as, age, gender, region, insurance type and race/ethnicity were assessed on index date. Baseline comorbidities were identified during the baseline period using ICD-10-CM diagnosis or procedure, HCPCS, and CPT-4 codes, and included as binary variables. Utilizations of other vaccinations or cancer screening during the baseline period was described. Immunocompromised status was derived based on previously established algorithm.^10,11^ The list of full variables with descriptive statistics is provided in supplemental table 3.

### Statistical methods

Categorical baseline characteristics were reported as frequency (n) and percentage. Continuous variables were reported as mean with standard deviation (SD) and median with interquartile. Absolute standardized differences (ASDs) were calculated to compare the distribution of baseline demographic and characteristics between the two cohorts. Hospitalization outcome was analyzed by the proportion of individuals with one or more hospitalization. OP/ER outcomes were analyzed by the mean number of outcomes.

We used the stabilized inverse probability of treatment weighting (IPTW) to adjust for potential confounding. Baseline demographic, clinical characteristics that were identified as potential confounders and had ASD >0.1 were included in the IPTW calculation. The list of selected baseline variables and their descriptive statistics are shown in Table 2. ASDs were calculated after weighting again to determine whether there is residual imbalance among the measured variables.

Generalized linear models (GLM) were applied to estimate the association between vaccination status and HCRU or cost outcomes during follow-up. For count-type OP/ER outcomes, Poisson distribution with log link function was used to estimate the rate ratio. Pearson Chi-square scale was used to adjust for overdispersion of count data. For right-skewed cost outcome, gamma distribution with log-link was used to estimate the cost ratio. A negligible cost was added (0.1 cent) to patients with zero cost. For binary-type hospitalization outcome, robust Poisson regression was applied to estimate the relative risk.^12^ All statistical analyses were performed with 2-sided alpha at 0.05 level. Data extraction and cleaning were conducted using the Aetion Evidence Platform. Statistical analyses were conducted using SAS® version 9.4 (SAS Institute Inc., NC, USA).

### Ethics review

The study dataset only contains de-identified data as per the de-identification standard defined in Section §164.514(a) of the Health Insurance Portability and Accountability Act of 1996 (HIPAA) Privacy Rule. The process by which the data is de-identified is attested to through a formal determination by a qualified expert as defined in Section §164.514(b)(1) of the HIPAA Privacy Rule. Therefore, the study was considered exempt from Institutional Review Board approval and for obtaining informed consent.

## Results

### Baseline demographics and clinical characteristics

Table 1 provides the sample attrition table. Total 1,953,167 individuals met the study inclusion and exclusion criteria. Based on their vaccination status, 613,156 and 1,340,011 individuals were included in the co-admin and influenza cohort respectively.

**Table 1:** Attrition table summarizing inclusion and exclusion criteria.

|  | **Co-admin cohort** | **Influenza cohort** |
| --- | --- | --- |
| Patients were identified from Optum CDM during Aug 31, 2022- Jul31, 2023. | Who had received COVID-19 and influenza vaccine on the same day: 1,425,838 | Who had received at least one influenza vaccine: 6,224,981 |
| **Exclusion** |  |  |
| 1. Insufficient enrollment during baseline and till July 31, 2023 | 904,331 | 3,957,199 |
| 2. Missing demographics (age, sex, payer, state) | 903,736 | 3,954,266 |
| 3. Age < 18 on index date | 856,825 | 3,606,716 |
| 4. Any COVID-19 and/or Influenza Vaccine during Aug1-30, 2022 | 847,668 | 3,540,628 |
| 5. For co-admin cohort: more than one COVID and/or Influenza vaccine during Aug 31, 2022 – Jul 31, 2023* | 777,928 | 1,655,246 |
| For influenza cohort: any COVID-19 vaccine or more than one influenza vaccine during Aug 31, 2022 - Jul31, 2023 |  |  |
| 6. Index vaccination after January 31, 2023 | 769,241 | 1,617,185 |
| 7. Patients under 50-year-old | 613,156 | 1,340,011 |
| **Final Cohort** | **613,156** | **1,340,011** |

\*For the co-admin cohort, 1086 individuals were removed as they received more than 2 COVID-19 vaccines. 58,554 individuals were removed as they received 2 COVID-19 vaccines.

Table 2 summarizes the selected baseline demographic and clinical characteristics for the two cohorts (Supplementary table 3 for the entire table). The mean age of the cohorts was 71 and 72 for co-admin and influenza cohort, respectively. Across both cohorts, most of the individuals were 65+ and hence were predominantly insured by Medicare Advantage plans. The overall comorbidity burden quantified by mean Charlson-Quan comorbidity index score was under 1 in both cohorts, though a trend towards influenza cohort having higher proportion of individuals with specific comorbidity was observed. In co-admin cohort during the baseline period, higher proportion of individuals utilized preventive health services such as other vaccines and screening procedures, while the level of healthcare utilization (hospitalization, OP/ER encounters) was lower compared to influenza cohort. All baseline variables were well balanced after weighting.

**Table 2:** Baseline demographic and clinical characteristics.

|  | **Co-admin Cohort**<br>**N = 613,156** | **Influenza Cohort**<br>**N =1,340,011** | **ASD (Prior**<br>**to**<br>**Weighting)** | **ASD (Post-**<br>**Weighting)** |
| --- | --- | --- | --- | --- |
| **Age on Index Date** |  |  |  |  |
| mean (SD) | 71 (9.18) | 72 (9.42) | 0.097 | 0.009 |
| median (IQR) | 71.00 (66.00,77.00) | 72.00<br>(67.00,79.00) |  |  |
| **Gender, n(%)** |  |  |  |  |
| Female | 327,738 (53.45%) | 775,100 (57.84%) | 0.088 | 0.003 |
| Male | 285,418 (46.55%) | 564,911 (42.16%) | 0.088 | 0.003 |
| **Race, n(%)** |  |  |  |  |
| White | 476,306 (77.68%) | 948,828 (70.81%) | 0.158 | 0.021 |
| Asian | 20,011 (3.26%) | 45,918 (3.43%) | 0.009 | 0.008 |
| Black | 44,449 (7.25%) | 133,074 (9.93%) | 0.096 | 0.005 |
| Hispanic | 39,092 (6.38%) | 126,389 (9.43%) | 0.113 | 0.02 |
| Missing/unknown | 33,298 (5.43%) | 85,802 (6.40%) | 0.041 | 0.003 |
| **Insurance, n(%)** |  |  |  |  |
| Commercial | 125,618 (20.49%) | 222,061 (16.57%) | 0.101 | 0.014 |
| Medicare<br>Advantage | 487,538 (79.51%) | 1,117,950 (83.43%) | 0.101 | 0.014 |
| **Region, n(%)** |  |  |  |  |
| Northeast | 76,274 (12.44%) | 169,717 (12.67%) | 0.007 | 0.002 |
| Midwest | 169,838 (27.70%) | 279,674 (20.87%) | 0.16 | 0.001 |
| South | 197,075 (32.14%) | 636,165 (47.47%) | 0.317 | 0.001 |
| West | 169,969 (27.72%) | 254,455 (18.99%) | 0.207 | 0.002 |
| **Charlson-Quan score** |  |  |  |  |
| mean (SD) | 0.40 (0.86) | 0.51 (0.94) | 0.121 | 0.004 |
| median (IQR) | 0.00 (0.00,1.00) | 0.00 (0.00,1.00) |  |  |
| **Comorbidities, n(%)** |  |  |  |  |
| Anemia | 87,415 (14.26%) | 245,452 (18.32%) | 0.11 | 0.003 |
| Asthma | 42,702 (6.96%) | 101,029 (7.54%) | 0.022 | 0.001 |
| Chronic kidney<br>disease | 101,050 (16.48%) | 271,729 (20.28%) | 0.098 | 0.002 |
| Hypertension | 385,012 (62.79%) | 945,512 (70.56%) | 0.165 | 0.003 |
| Obesity | 121,794 (19.86%) | 276,265 (20.62%) | 0.019 | 0.003 |
| Influenza infection | 2,887 (0.47%) | 10,386 (0.78%) | 0.039 | 0.003 |
| Smoking | 107,972 (17.61%) | 255,674 (19.08%) | 0.038 | 0.003 |
| COVID-19 Infection | 54,125 (8.83%) | 169,947 (12.68%) | 0.125 | 0.004 |
| Pneumonia | 15,499 (2.53%) | 53,782 (4.01%) | 0.084 | 0.004 |
| **Receipt of COVID-19 vaccine in prior year, n(%)** | 366,183 (59.72%) | 373,455 (27.87%) | 0.678 | 0.007 |
| **Receipt of influenza vaccine in prior year, n (%)** | 259,654 (42.35%) | 495,290 (36.96%) | 0.11 | 0.012 |
| **Other Vaccine Utilization, n(%)** | 126,327 (20.60%) | 222,027 (16.57%) | 0.104 | 0.004 |
| **Any Cancer screening procedure, n(%)** | 235,000 (38.33%) | 488,070 (36.42%) | 0.039 | 0.006 |
| **Immunocompromised status, n(%)** | 80,046 (13.05%) | 185,017 (13.81%) | 0.022 | 0.002 |
| **Hospitalizations during baseline, n(%)** | 47,609 (7.76%) | 139,409 (10.40%) | 0.092 | 0.005 |
| **Number of all-cause OP/ER encounter at baseline** |  |  |  |  |
| mean (SD) | 17.92 (18.75) | 20.60 (23.46) | 0.126 | 0.001 |
| median (IQR) | 12.00 (6.00,23.00) | 14.00 (7.00,26.00) |  |  |
| **Index Vaccination Year and Month, n(%)** |  |  |  |  |
| 2022 AUG-SEP | 165,517 (26.99%) | 277,656 (20.72%) | 0.148 | 0.015 |
| 2022 OCT | 286,133 (46.67%) | 579,235 (43.23%) | 0.069 | 0.008 |
| 2022 NOV | 105,510 (17.21%) | 303,245 (22.63%) | 0.136 | 0.009 |
| 2022 DEC | 43,530 (7.10%) | 133,756 (9.98%) | 0.103 | 0.016 |
| 2023 JAN | 12,466 (2.03%) | 46,119 (3.44%) | 0.086 | 0.008 |

SD: standard deviation; IQR: interquartile range; ASD: absolute standardized difference; OP/ER: outpatient or emergency room

### HCRU and cost outcomes

The co-administration of influenza and COVID-19 vaccines on the same day was associated with 5% lower risk of all-cause hospitalizations (RR: 0.95, 95%CI: 0.93-0.96), 41% lower risk of COVID-19-related hospitalizations (RR: 0.59, 95%CI: 0.56-0.63), 6% lower risk of cardiorespiratory-related hospitalizations (RR: 0.94, 95%CI: 0.93-0.96), and 14% lower risk of pneumonia-related hospitalizations (RR: 0.86, 95%CI: 0.83-0.90), compared to receiving influenza vaccine only. The association between vaccination status and influenza-related hospitalizations was not statistically significant (RR: 0.91, 95%CI: 0.81, 1.04). Similar trends were also observed for number of OP/ER encounters. (Table 3).

**Table 3:** Association between vaccination status and HCRU.

| outcomes | Co-admin<br>Cohort | Influenza<br>Cohort | Weighted RR<br>(95% CI)** |
| --- | --- | --- | --- |
| **All-cause** |  |  |  |
| Hospitalizations during follow-up, (95% CI)* | 5.19% (5.13, 5.24) | 6.65% (6.61, 6.69) | 0.95 (0.93, 0.96) |
| Mean number of OP/ER encounter during follow-up, (95% CI)# | 9.204 (9.177, 9.230) | 10.800 (10.777, 10.823) | 0.96 (0.95, 0.96) |
| **COVID-19-related** |  |  |  |
| Hospitalizations during follow-up, (95% CI)* | 0.27% (0.26, 0.28) | 0.53% (0.52, 0.54) | 0.59 (0.56, 0.63) |
| Mean number of OP/ER encounter during follow-up, (95% CI)# | 0.067 (0.065, 0.069) | 0.110 (0.109, 0.112) | 0.68 (0.66, 0.70) |
| **Cardiorespiratory-related** |  |  |  |
| Hospitalizations during follow-up, (95% CI)* | 4.62% (4.57, 4.68) | 6.07% (6.03, 6.11) | 0.94 (0.93, 0.96) |
| Mean number of OP/ER encounter during follow-up, (95% CI)# | 3.125 (3.110, 3.140) | 4.125 (4.113, 4.138) | 0.94 (0.93, 0.94) |
| **Pneumonia-related** |  |  |  |
| Hospitalizations during follow-up, (95% CI)* | 0.71% (0.69, 0.73) | 1.09% (1.07, 1.11) | 0.86 (0.83, 0.90) |
| Mean number of OP/ER encounter during follow-up, (95% CI)# | 0.059 (0.057, 0.061) | 0.094 (0.092, 0.096) | 0.87(0.84, 0.91) |
| **Influenza-related** |  |  |  |
| Hospitalizations during follow-up, (95% CI)* | 0.07% (0.07, 0.08) | 0.10% (0.09, 0.10) | 0.91 (0.81, 1.04) |
| Mean number of OP/ER encounter during follow-up, (95% CI)# | 0.012 (0.012, 0.013) | 0.017 (0.016, 0.017) | 0.86 (0.81, 0.92) |

\*\*influenza cohort as the reference
\* Relative risk is calculated by robust Poisson regression

All-cause medial costs during the follow-up period for two cohorts are summarized in Table 4. On average, individuals in co-admin cohort had lower cost ($7,868) than individuals in influenza cohort ($9,415). After adjusting for baseline variables, co-admin cohort had 3% less all-cause medical cost (cost ratio = 0.974, 95% CI: 0.968, 0.979) during the follow-up period compared to influenza cohort.

**Table 4:** Association between vaccination status and all-cause medical costs.

|  | **Co-admin** | **Influenza** | **Weighted cost ratio (95% CI)** |
| --- | --- | --- | --- |
| N* | 612,689 | 1,339,079 |  |
| Mean (SD) | \$7,868 (\$21,492) | \$9,415 (\$25,025) | 0.974 (0.968, 0.979) ** |
| Median (IQR) | \$1,853 (\$596, \$5,730) | \$2,200 (\$726, \$6,844) | |

\*Patients with negative cost during baseline or follow-up had been excluded.
\*\*IPTW was recalculated after exclusion. Generalized linear model with gamma distribution and log link function was used to estimate the cost ratio. Zero cost had been replaced with 1/10 cent.
SD: standard deviation; IQR: interquartile range

## Discussion

The study result indicates that receiving both influenza and COVID-19 vaccination on same day was associated with significantly reduced all-cause, COVID-19 related, pneumonia related, and cardiorespiratory related hospitalization risks and average number of OP/ER visits compared to receiving influenza vaccine only during the 2022/2023 season. The reduction in risk was more pronounced for COVID-19 related hospitalization. Driven by reduced hospitalization and OP/ER visits, all cause medical cost was significantly lower with vaccine co-administration on same day as compared to receiving influenza vaccine only. To our knowledge, this is among the first studies to compare medical costs associated with co-administration of COVID-19 and influenza vaccines.

We found that co-admin cohort had reduced risk of influenza-related OP/ER encounters than influenza cohort. Excluding the possibility of a chance finding, this observation can be a result of residual confounding given that influenza cohort in general had higher medical needs. McGrath et al. in a retrospective cohort study of US adults also concluded that among individuals 65 years or older, co-administration of BNT162b2 BA.4/5 bivalent mRNA COVID-19 vaccine and influenza vaccine was associated with lower incidence of influenza-related outcomes compared to receiving influenza vaccine alone. When they calibrated using negative control outcomes, the observed association moved closer to the null and was no longer statistically significant.^13^

Following favorable evidence from cohort study and clinical trials^14–16^ on safety and immunogenicity, co-administration of influenza and COVID-19 vaccines on the same day have been recommended by most public authorities,^17^ including the CDC.^5^ The safety of co-administration strategy was further studied in a recent VAERS database analysis which concluded that co-administration of bivalent mRNA COVID-19 and seasonal influenza vaccines did not reveal any unusual or unexpected patterns of AEs.^18^ CDC ACIP recently proposed changing the COVID-19 recommendation timelines from current Sept to June to align with the influenza recommendation timelines.^19^ However, the uptake of co-administration strategy is still growing. Among a large sample of older U.S. Medicare beneficiaries who received an mRNA COVID-19 booster vaccine, the prevalence of vaccine co-administration with an influenza vaccine was 11.1% in 2021 and 36.5% in 2022, and varied widely across U.S. counties.^20^

Various aspects of the benefits with both influenza and COVID-19 vaccination have been described in prior literature. Recent cost effectiveness analyses by CDC concludes that influenza vaccination results in net health gains but also incurs net costs for all adult age cohorts, except for vaccination of high-risk adults >50 years which was projected to be cost saving.^21^ Similarly, vaccination against COVID-19 has been shown to be cost effective and reduce burden to the health care system.^22^ Ensuring sufficient vaccine coverage for both COVID-19 and influenza is a crucial driver to reduce the burden to health care system of significant respiratory illnesses. Improvement in vaccination coverage and likelihood of completing all recommended vaccinations through use of a multi-component, combination vaccine was previously demonstrated in pediatric space.^23,24^ Several multi-component combination vaccines against influenza and COVID-19 are in clinical development,^25–27^ which if approved could help in increasing vaccine coverage and reduce avoidable burden to health care system.

This study has several strengths worth highlighting. It was conducted on a large insurance claims database including both commercially and Medicare Advantage insured population. This study leveraged contemporary data from the most recent complete season during which the US COVID-19 was in an endemic setting. We were able to account for a comprehensive list of baseline variables to adjust for confounding using the stabilized IPTW, including preventive healthcare utilizations that could approximate the unmeasured health seeking behavior.

There are a few limitations to this study. First, given the nature of observational study, there is a possibility of residual confounding bias. Even though we controlled for a large number of baseline variables, unmeasured confounders such as occupation, lifestyle, or social interaction pattern can exist. Second, all variables, including exposure, outcomes, and baseline covariates, were derived from claims data and are susceptible to measurement errors. Such errors in this study are most likely to be non-differential between two cohorts and would bias the result towards null. Third, we imposed the continuous enrollment criteria for both baseline and follow-up period, which can introduce selection bias. Thus, the association observed in this study should be interpreted carefully and extrapolating results to other settings in terms of population, season, or age groups should be cautiously done. The current analyses were only conducted over one season and would benefit from updating or replicating with more recent season data in the future. The analyses did not consider individuals who received both influenza and COVID-19 vaccines but on separate days. The association for this type of vaccination pattern with outcomes is a topic for future research.

## Conclusion

Results from the study suggests that receiving both COVID-19 and influenza vaccines on the same day in comparison to receipt of influenza vaccines only was associated with reduced downstream utilization of healthcare resources, especially the risk of COVID-19 related hospitalization and all-cause medical costs. Thus, interventions that could help increase vaccine coverage, especially for COVID-19 might have economic and public health benefits.

## Supporting information

Mehta Supplement medRxiv 2024

## Data Availability

The analytical dataset used in the analyses would be made available at reasonable request.

## Notes

### Competing Interest Statement

All authors are employees of Moderna Inc. and might own stocks in the organization

### Funding Statement

The study was funded by Moderna, Inc.

