## Supplementary material for "Comparison of healthcare resource use and cost between influenza and COVID-19 vaccine coadministration and influenza vaccination only": Mehta Supplement medRxiv 2024

**Supplementary Table 1: Procedure and NDC codes used to identify COVID-19 and influenza vaccinations**

| Vaccine | Code Type | Code |
|---|---|---|
| Influenza vaccine | HCPCS/CPT | 90630, 90653, 90654, 90655, 90656, 90657, 90658, 90660, 90661, 90662, 90664, 90666, 90667, 90668, 90672, 90673, 90674, 90682, 90685, 90686, 90687, 90688, 90689, 90694, 90756, G0008, Q2034, Q2035, Q2036, Q2037, Q2038, Q2039 |
| | NDC | 19515080852, 19515081652, 19515081852, 19515084511, 19515088907, 19515089007, 19515089111, 19515089307, 19515089452, 19515089511, 19515089611, 19515089711, 19515089811, 19515090011, 19515090152, 19515090311, 19515090652, 19515090852, 19515090952, 19515091252, 33332001001, 33332001301, 33332001401, 33332001501, 33332001601, 33332001701, 33332001801, 33332011010, 33332011310, 33332011410, 33332011510, 33332011610, 33332011710, 33332011810, 33332021920, 33332022020, 33332022120, 33332031601, 33332031701, 33332031801, 33332031901, 33332032001, 33332032101, 33332032203, 33332041610, 33332041710, 33332041810, 33332041910, 33332042010, 33332042110, 33332042210, 42874001210, 42874001310, 42874001410, 42874001510, 42874001610, 42874001710, 42874011710, 49281001010, 49281001025, 49281001050, 49281001110, 49281001150, 49281001210, 49281001250, 49281001310, 49281001350, 49281001450, 49281011125, 49281011225, 49281011325, 49281012065, 49281012165, 49281012265, 49281032050, 49281032150, 49281033615, 49281033715, 49281038615, 49281038765, 49281038815, 49281038965, 49281039015, 49281039165, 49281039215, 49281039365, 49281039415, 49281039565, 49281039615, 49281039765, 49281039965, 49281040165, |

| | | |
|---|---|---|
| | | 49281040365, 49281040565, 49281041310, 49281041350, 49281041410, 49281041450, 49281041510, 49281041610, 49281041650, 49281041710, 49281041750, 49281041810, 49281041850, 49281041910, 49281041950, 49281042010, 49281042050, 49281042110, 49281042150, 49281042210, 49281042250, 49281051325, 49281051425, 49281051625, 49281051725, 49281051825, 49281051925, 49281052025, 49281052125, 49281062115, 49281062515, 49281062715, 49281062915, 49281063115, 49281063315, 49281063515, 49281063715, 49281070355, 49281070555, 49281070755, 49281070840, 49281070955, 49281071040, 49281071240, 49281071810, 49281071910, 49281072010, 49281072110, 49281072210, 54868617700, 54868618000, 58160087952, 58160088052, 58160088152, 58160088352, 58160088552, 58160088752, 58160089052, 58160089652, 58160089852, 58160090052, 58160090152, 58160090352, 58160090552, 58160090752, 62577061301, 62577061401, 63851061201, 66019010701, 66019010810, 66019010910, 66019011010, 66019030010, 66019030110, 66019030210, 66019030310, 66019030410, 66019030510, 66019030610, 66019030710, 66019030810, 66019030910, 66521000001, 66521011202, 66521011210, 66521011302, 66521011310, 66521011402, 66521011410, 66521011502, 66521011510, 66521011602, 66521011610, 66521011702, 66521011710, 66521011802, 66521011810, 70461000101, 70461000201, 70461001803, 70461001903, 70461002003, 70461011902, 70461011910, 70461012002, 70461012003, 70461012010, 70461012103, 70461012203, 70461020001, 70461020101, 70461030110, 70461031803, 70461031903, 70461032003, 70461032103, 70461032203, 70461041810, 70461041910, 70461042010, |

| | | |
|---|---|---|
| | | 70461042110, 70461042210, 76420048201, 76420048301, 49281018125 |
| COVID-19 - vaccine | HCPCS/CPT | 0021A, 0022A, 91302, 0031A, 0034A, 91303, 0011A, 0012A, 0013A, 0064A, 0091A, 0092A, 0093A, 0094A, 0111A, 0112A, 0113A, 0134A, 0141A, 0142A, 0144A, 0164A, 91301, 91306, 91309, 91311, 91313, 91314, 91316, 91321, 91322, 0041A, 0042A, 0044A, 91304, 90480, 0001A, 0002A, 0003A, 0004A, 0051A, 0052A, 0053A, 0054A, 0071A, 0072A, 0073A, 0074A, 0081A, 0082A, 0083A, 0121A, 0124A, 0151A, 0154A, 0171A, 0172A, 0173A, 0174A, 91300, 91305, 91307, 91308, 91312, 91315, 91317, 91318, 91319, 91320, 0104A, 91310 |
| | NDC | 00310122210, 00310122215, 0310122210, 0310122215, 59676058005, 59676058015, 5967658005, 5967658015, |

| | | |
|---|---|---|
| | | 80777010011, 80777010099, 80777010201, 80777010204, 80777010293, 80777010295, 80777010296, 80777027310, 80777027315, 80777027398, 80777027399, 80777027505, 80777027599, 80777027705, 80777027799, 80777027905, 80777027999, 80777028005, 80777028099, 80777028205, 80777028299, 80777028302, 80777028399, 80777028707, 80777028792, 8077701001, 8077710011, 8077710099, 8077710201, 8077710204, 8077710293, 8077710295, 8077710296, 8077727310, 8077727315, 8077727398, 8077727399, 8077727505, 8077727599, 8077727705, 8077727799, 8077727905, 8077727999, 8077728205, 8077728299, 8077728302, 8077728399, 8077728707, 8077728792, 80631010001, 80631010010, 80631010501, 80631010502, 80631100001, 8063110001, 8063110010, 8063110201, 8063110210, 8063110501, 8063110502, 80631010201, 80631010210, 00069202501, 00069202510, 00069202525, 00069236201, 00069236210, 00069239201, 00069239210, 59267007801, 59267007804, 59267030401, 59267030402, 59267056501, 59267056502, 59267060901, 59267060902, 59267100001, 59267100002, 59267100003, 59267102501, 59267102502, 59267102503, 59267102504, 59267105501, 59267105502, 59267105504, 59267140401, 59267140402, 59267431502, 59267433101, 59267433102, 0006920251, 5926700781, 5926700784, 5926703041, 5926703042, 5926705651, 5926705652, 5926706091, 5926706092, 5926710001, 5926710002, 5926710003, 5926710251, 5926710252, 5926710253, 5926710254, 5926710551, 5926710552, 5926710554, 5926714041, 5926714042, 5926743151, 5926743152, 5926743311, 5926743312, 49281061878, 49281061820, 4928161820, 4928161878 |

**Supplementary Table 2: International Classification of Diseases, 10th Revision (ICD-10) diagnostic codes used to identify pneumonia, influenza, COVID-19 and cardiorespiratory conditions**

| **Condition** | **Code** |
|---|---|
| Pneumonia | J12, J12.0, J12.09, J12.1, J12.10, J12.2, J12.3, J12.59, J12.8, J12.81, J12.82, J12.84, J12.89, J12.9, J12.9X, J13, J13.0, J13.09, J13.2, J13.35, J13.9, J14, J14.3, J14.8, J14.9, J15, J15.0, J15.02XA, J15.1, J15.2, J15.20, J15.21, J15.211, J15.212, J15.29, J15.3, J15.4, J15.5, J15.6, J15.61, J15.69, J15.7, J15.8, J15.82, J15.9, J16, J16.0, J16.8, J16.80, J16.9, J17, J17.0, J17.210, J17.8, J18, J18.0, J18.00, J18.1, J18.10, J18.18, J18.2, J18.20, J18.3, J18.6, J18.7, J18.8, J18.80, J18.85, J18.89, J18.9, J18.90, J18.9000, J18.91, J18.96, J18.97, J18.9D |
| Influenza | J09, J09.0, J09.02, J09.10, J09.2, J09.2X, J09.6, J09.7X, J09.81, J09.82, J09.9, J09.90, J09.X, J09.X1, J09.X2, J09.X3, J09.X9, J09.XA, J09.XI, J10, J10.0, J10.00, J10.01, J10.08, J10.1, J10.10, J10.110, J10.12, J10.17, J10.19, J10.190, J10.2, J10.3, J10.4, J10.40, J10.45, J10.8, J10.80, J10.81, J10.82, J10.83, J10.89, J10.9, J10.90, J11, J11.0, J11.00, J11.08, J11.1, J11.10, J11.103, J11.11, J11.2, J11.20, J11.23, J11.5, J11.8, J11.81, J11.82, J11.83, J11.89, J11.9 |
| COVID-19 | U07.1 |
| Cardiorespiratory | I**, J** |

Supplemental Table 3

| | **Co-admin N = 613,156** | **Influenza N =1,340,011** | **ASD (Prior to Weighting)** | **ASD (Post-Weighting)** |
|---|---|---|---|---|
| **Age on Index Date** | | | | |
| mean (SD) | 70.97 (9.18) | 71.87 (9.42) | 0.097 | 0.009 |
| median (IQR) | 71.00 (66.00,77.00) | 72.00 (67.00,79.00) | | |
| **Age Group, n(%)** | | | | |
| 50-64 | 132,848 (21.67%) | 268,217 (20.02%) | 0.041 | 0.023 |
| 65+ | 480,308 (78.33%) | 1,071,794 (79.98%) | 0.041 | 0.023 |
| **Gender, n(%)** | | | | |
| Female | 327,738 (53.45%) | 775,100 (57.84%) | 0.088 | 0.003 |
| Male | 285,418 (46.55%) | 564,911 (42.16%) | 0.088 | 0.003 |
| **Race, n(%)** | | | | |
| White | 476,306 (77.68%) | 948,828 (70.81%) | 0.158 | 0.021 |
| Asian | 20,011 (3.26%) | 45,918 (3.43%) | 0.009 | 0.008 |
| Black | 44,449 (7.25%) | 133,074 (9.93%) | 0.096 | 0.005 |
| Hispanic | 39,092 (6.38%) | 126,389 (9.43%) | 0.113 | 0.02 |
| Missing/unknown | 33,298 (5.43%) | 85,802 (6.40%) | 0.041 | 0.003 |
| **Insurance, n(%)** | | | | |
| Commercial | 125,618 (20.49%) | 222,061 (16.57%) | 0.101 | 0.014 |
| Medicare Advantage | 487,538 (79.51%) | 1,117,950 (83.43%) | 0.101 | 0.014 |
| **Region, n(%)** | | | | |
| Northeast | 76,274 (12.44%) | 169,717 (12.67%) | 0.007 | 0.002 |
| Midwest | 169,838 (27.70%) | 279,674 (20.87%) | 0.16 | 0.001 |
| South | 197,075 (32.14%) | 636,165 (47.47%) | 0.317 | 0.001 |
| West | 169,969 (27.72%) | 254,455 (18.99%) | 0.207 | 0.002 |
| **Charlson-Quan score, n(%)** | | | | |

| | | | | |
|---|---|---|---|---|
| 0 | 448,619 (73.17%) | 898,530 (67.05%) | 0.134 | 0.015 |
| 1 | 113,722 (18.55%) | 289,525 (21.61%) | 0.076 | 0.017 |
| >=2 | 50,815 (8.29%) | 151,956 (11.34%) | 0.103 | 0.001 |
| **Charlson-Quan score** | | | | |
| mean (SD) | 0.40 (0.86) | 0.51 (0.94) | 0.121 | 0.004 |
| median (IQR) | 0.00 (0.00,1.00) | 0.00 (0.00,1.00) | | |
| **Comorbidities, n(%)** | | | | |
| Alcohol Abuse | 1,107 (0.18%) | 2,203 (0.16%) | 0.004 | 0.007 |
| Anemia | 87,415 (14.26%) | 245,452 (18.32%) | 0.11 | 0.003 |
| Asthma | 42,702 (6.96%) | 101,029 (7.54%) | 0.022 | 0.001 |
| Cerebrovascular Disease | 51,833 (8.45%) | 146,653 (10.94%) | 0.084 | 0.011 |
| Chronic Pulmonary Disease | 100,103 (16.33%) | 262,332 (19.58%) | 0.085 | 0.025 |
| Chronic kidney disease | 101,050 (16.48%) | 271,729 (20.28%) | 0.098 | 0.002 |
| Coagulopathy | 35,311 (5.76%) | 92,148 (6.88%) | 0.046 | 0.018 |
| Congestive Heart Failure | 51,529 (8.40%) | 148,251 (11.06%) | 0.09 | 0.006 |
| COPD | 65,744 (10.72%) | 187,197 (13.97%) | 0.099 | 0.033 |
| Cystic Fibrosis | 58 (0.01%) | 126 (0.01%) | 0 | 0.002 |
| Dementia | 18,029 (2.94%) | 55,039 (4.11%) | 0.063 | 0.002 |
| Depression | 95,994 (15.66%) | 215,175 (16.06%) | 0.011 | 0.037 |
| Dysautonomia | 445 (0.07%) | 958 (0.07%) | 0 | 0.007 |
| Fluid and Electrolyte Disorders | 52,674 (8.59%) | 152,025 (11.35%) | 0.092 | 0.008 |
| Hypertension | 385,012 (62.79%) | 945,512 (70.56%) | 0.165 | 0.003 |
| Hyperthyroidism | 5,708 (0.93%) | 15,114 (1.13%) | 0.02 | 0.006 |
| Liver Disease | 34,418 (5.61%) | 88,710 (6.62%) | 0.042 | 0.003 |

| | | | | |
|---|---|---|---|---|
| Myocardial Infarction | 20,532 (3.35%) | 57,984 (4.33%) | 0.051 | 0.007 |
| Obesity | 121,794 (19.86%) | 276,265 (20.62%) | 0.019 | 0.003 |
| Neurological Disorders | 35,810 (5.84%) | 91,981 (6.86%) | 0.042 | 0.008 |
| Influenza infection | 2,887 (0.47%) | 10,386 (0.78%) | 0.039 | 0.003 |
| RSV infection | 409 (0.07%) | 1,226 (0.09%) | 0.009 | 0 |
| Paralysis | 4,092 (0.67%) | 12,196 (0.91%) | 0.027 | 0.002 |
| Peptic Ulcer Disease | 5,713 (0.93%) | 14,973 (1.12%) | 0.018 | 0.008 |
| Peripheral Vascular Disease | 91,914 (14.99%) | 254,247 (18.97%) | 0.106 | 0.026 |
| Postural orthostatic tachycardia syndrome | 6,036 (0.98%) | 15,324 (1.14%) | 0.016 | 0.002 |
| Psychoses | 3,441 (0.56%) | 10,467 (0.78%) | 0.027 | 0.009 |
| Pulmonary circulation disorder | 19,272 (3.14%) | 51,712 (3.86%) | 0.039 | 0.004 |
| Pulmonary fibrosis | 6,723 (1.10%) | 18,891 (1.41%) | 0.028 | 0.008 |
| Renal Failure | 94,863 (15.47%) | 254,569 (19.00%) | 0.093 | 0 |
| Rheumatoid Arthritis | 15,594 (2.54%) | 42,753 (3.19%) | 0.039 | 0.003 |
| Sickle cell disease | 321 (0.05%) | 1,000 (0.07%) | 0.009 | 0.002 |
| Smoking | 107,972 (17.61%) | 255,674 (19.08%) | 0.038 | 0.003 |
| Type 1 Diabetes | 6,601 (1.08%) | 17,634 (1.32%) | 0.022 | 0.002 |
| Type 2 Diabetes | 156,829 (25.58%) | 425,649 (31.76%) | 0.137 | 0.041 |
| Valvular Disease | 62,783 (10.24%) | 165,680 (12.36%) | 0.067 | 0.008 |
| COVID-19 Infection | 54,125 (8.83%) | 169,947 (12.68%) | 0.125 | 0.004 |

| | | | | |
|---|---|---|---|---|
| Post COVID-19 Condition | 2,882 (0.47%) | 10,078 (0.75%) | 0.036 | 0.01 |
| Pneumonia | 15,499 (2.53%) | 53,782 (4.01%) | 0.084 | 0.004 |
| **Receipt of COVID-19 vaccine in prior year, n(%)** | 366,183 (59.72%) | 373,455 (27.87%) | 0.678 | 0.007 |
| **Receipt of influenza vaccine in prior year, n (%)** | 259,654 (42.35%) | 495,290 (36.96%) | 0.11 | 0.012 |
| **Other Vaccine Utilization, n(%)** | 126,327 (20.60%) | 222,027 (16.57%) | 0.104 | 0.004 |
| Haemophilus influenzae type b Vaccine | 118 (0.02%) | 229 (0.02%) | 0.002 | 0.001 |
| Hepatitis A Vaccine | 854 (0.14%) | 1,065 (0.08%) | 0.018 | 0.007 |
| Hepatitis B Vaccine | 3,002 (0.49%) | 7,065 (0.53%) | 0.005 | 0.013 |
| Human papillomavirus (HPV) Vaccine | 58 (0.01%) | 69 (0.01%) | 0.005 | 0.002 |
| Measles, mumps, rubella (MMR) - Vaccine | 298 (0.05%) | 417 (0.03%) | 0.009 | 0.001 |
| Meningococcal Vaccine | 482 (0.08%) | 515 (0.04%) | 0.017 | 0.011 |
| Pneumococcal Vaccine | 43,883 (7.16%) | 84,364 (6.30%) | 0.034 | 0.01 |

| | | | | |
|---|---|---|---|---|
| Tetanus, diphtheria, pertussis (TDaP) Vaccine | 35,040 (5.71%) | 60,330 (4.50%) | 0.055 | 0.003 |
| Varicella Vaccine | 47 (0.01%) | 62 (0.00%) | 0.004 | 0.001 |
| Zoster Vaccine | 62,434 (10.18%) | 98,163 (7.33%) | 0.101 | 0.011 |
| **Any Cancer screening procedure, n(%)** | 235,000 (38.33%) | 488,070 (36.42%) | 0.039 | 0.006 |
| Breast cancer screening procedure | 143,920 (23.47%) | 306,794 (22.89%) | 0.014 | 0.002 |
| Cervical cancer screening procedure | 10,400 (1.70%) | 21,681 (1.62%) | 0.006 | 0.007 |
| Colorectal cancer screening procedure | 68,844 (11.23%) | 139,847 (10.44%) | 0.025 | 0.002 |
| Prostate cancer screening procedure | 48,178 (7.86%) | 93,589 (6.98%) | 0.033 | 0.005 |
| **Immunocompromised status, n (%)** | 80,046 (13.05%) | 185,017 (13.81%) | 0.022 | 0.002 |
| Stem Cell Transplant during baseline | 5 (0.00%) | 39 (0.00%) | 0.005 | 0.004 |
| Blood Transplant within 730 days | 11,925 (1.94%) | 34,047 (2.54%) | 0.04 | 0.013 |
| Malignancy within 180 days | 51,870 (8.46%) | 117,724 (8.79%) | 0.012 | 0.002 |
| Ever HIV | 2,722 (0.44%) | 4,273 (0.32%) | 0.02 | 0.033 |
| Immunosuppressive Therapy within 60 days | 18,444 (3.01%) | 40,140 (3.00%) | 0.001 | 0.009 |
| Ever Primary Immunodeficiency | 3,397 (0.55%) | 8,317 (0.62%) | 0.009 | 0.005 |

| | | | | |
|---|---|---|---|---|
| Organ Transplant and Immunosuppressive Therapy within 60 days | 3,967 (0.65%) | 8,779 (0.66%) | 0.001 | 0.01 |
| **Hospitalizations during baseline, n (%)** | 47,609 (7.76%) | 139,409 (10.40%) | 0.092 | 0.005 |
| **Number of all-cause outpatient or ER encounter at baseline** | | | | |
| mean (SD) | 17.92 (18.75) | 20.60 (23.46) | 0.126 | 0.001 |
| median (IQR) | 12.00 (6.00,23.00) | 14.00 (7.00,26.00) | | |
| **Baseline all-cause medical cost ($) #** | | | | |
| mean (SD) | 13,945.37 (31,878.80) | 16,583.35 (40,920.78) | 0.072 | 0.017 |
| median (IQR) | 4,403.94 (1,597.90,12,015.46) | 5,130.68 (1,877.38,14,050.31) | | |
| **Index Year and Month, n(%)** | | | | |
| 2022 AUG-SEP | 165,517 (26.99%) | 277,656 (20.72%) | 0.148 | 0.015 |
| 2022 OCT | 286,133 (46.67%) | 579,235 (43.23%) | 0.069 | 0.008 |
| 2022 NOV | 105,510 (17.21%) | 303,245 (22.63%) | 0.136 | 0.009 |
| 2022 DEC | 43,530 (7.10%) | 133,756 (9.98%) | 0.103 | 0.016 |
| 2023 JAN | 12,466 (2.03%) | 46,119 (3.44%) | 0.086 | 0.008 |
